# Proteomic predictors of physical, cognitive and imaging outcomes in multiple sclerosis: 5-year follow-up study

**DOI:** 10.1101/2023.05.24.23290483

**Authors:** Kian Jalaleddini, Dejan Jakimovski, Anisha Keshavan, Shannon McCurdy, Kelly Leyden, Ferhan Qureshi, Atiyeh Ghoreyshi, Niels Bergsland, Michael G. Dwyer, Murali Ramanathan, Bianca Weinstock-Guttman, Ralph HB Benedict, Robert Zivadinov

## Abstract

**Background:** A quantitative measurement of serum proteome biomarkers that would associate with disease progression endpoints can provide risk stratification for persons with multiple sclerosis and supplement the clinical decision-making process.

**Materials and Methods:** 202 persons with multiple sclerosis were enrolled in a longitudinal study with measurements at two time points with an average follow-up time of 5.4 years. Clinical measures included the Expanded Disability Status Scale, Timed 25-foot Walk, 9-Hole Peg and Symbol Digit Modalities Tests. Subjects underwent magnetic resonance imaging to determine the volumetric measures of the whole brain, gray matter, deep gray matter and lateral ventricles. Serum samples were analyzed using a custom immunoassay panel on the Olink™ platform and concentrations of 18 protein biomarkers were measured. Linear mixed-effects models and adjustment for multiple comparisons were performed.

**Results:** Subjects had a significant 55.6% increase in Chemokine Ligand 20 (9.7pg/mL vs. 15.1pg/mL, p<0.001) and Neurofilament light polypeptide (10.5 pg/ml vs. 11.5 pg/ml, p=0.003) at the follow-up time point. Additional changes in CUB domain-containing protein 1, Contactin 2, Glial fibrillary acidic protein, Myelin oligodendrocyte glycoprotein, and Osteopontin were noted but did not survive multiple comparisons correction. Worse clinical performance in the 9-HPT was associated with Neurofilament light polypeptide (p=0.001). Increases in several biomarker candidates were correlated with greater neurodegenerative changes as measured by different brain volumes.

**Conclusion:** Multiple proteins representing diverse biological pathways (neuroinflammation, immune modulation, and neuroaxonal integrity) associate with physical, cognitive and radiographic outcomes. Future studies should determine the utility of multiple protein assays in routine clinical care.

## Introduction

Multiple sclerosis (MS) is a chronic, inflammatory, demyelinating, and degenerative disease of the central nervous system (CNS) resulting in progressive accrual of physical and cognitive disability.^1^ People with multiple sclerosis (pwMS) have heterogeneous clinical presentation and multiple demographics, clinical, and paraclinical risk factors have been associated with poorer long-term outcomes.^1^ Over the last three decades, a plethora of highly effective disease modifying therapies (DMTs) have been developed that can significantly lessen the long-term disability.^1^ They generally range from highly effective but immunosuppressive therapies to moderately effective immunomodulators with lower rate of adverse events.^2^ Therefore, development of attainable and cost-effective biomarkers can provide risk stratification for pwMS and supplement the clinical decision-making process.

In addition to validated imaging biomarkers such as lesion pathology and whole brain atrophy, recent developments in proteomic assay technology have allowed detection and measurement of picomolar concentrations of blood biomarkers.^3^ These biomarkers can provide proxy measures regarding the occurrence and extent of pathological changes in the CNS.^4^ For example, higher blood levels of neurofilament light chain (NfL), an intermediate filament present in neurons, can indicate greater neuroaxonal destruction, and higher levels of glial fibrillary acidic protein (GFAP) can indicate glial cell activation both indicative of presence of disease activity.^5, 6^ Although these individual biomarkers have already emerged as candidate outcomes measures in MS, a single protein biomarker can have limited ability in capturing changes within multiple parallel pathophysiological MS pathways.^7^ When compared to MRI measures, blood-derived analyses are less costly, more accessible, and can be bundled together with routine clinical blood work.^8^

The Multiple Sclerosis Disease Activity (MSDA) is a recently developed and analytically validated ^9^ panel of 18 biomarkers that represent changes within four main pathophysiological pathways of neuroinflammation, immunomodulation, myelin biology and neuroaxonal integrity.^9^ In the first clinical validation study, the MSDA platform was trained and tested as a predictor for presence of gadolinium-enhancing lesions or new/newly enlarging T2 lesions in a cohort of 614 samples.^10^ The multi-protein scores outperformed the best individual protein (NfL) with area under curve change from 0.726 to 0.781.^10^ Determining the relation of the MSDA panel to long-term disability outcomes and examining the longitudinal predictive properties of such an assay are essential for clinical adoption and wide-spread clinical utility.

The aims of this study were to determine the relationship between the multiple proteomic biomarkers and cross-sectional and longitudinal MS outcomes, including physical disability, cognitive performance and conventional MRI outcomes. We hypothesize that proteomic analysis could improve the clinical and MRI-based risk stratification when utilized in a heterogenous group of pwMS.

## Methods

### Study population

A total of 202 patients were assessed in these analyses and derived from a larger longitudinal, case-controlled study to explore the role of cardiovascular, environmental and genetic risk factors in multiple sclerosis patients (CEG-MS).^11^ In particular, for this study, the pwMS were enrolled at baseline between 2009-2012 and returned for a follow-up visit in years 2014-2017. The inclusion criteria were: 1) baseline age of 18-75 years old; 2) diagnosed with either MS or clinically isolated syndrome (CIS), defined by the 2010-revised McDonald criteria^12^; 3) availability of either baseline or follow-up serum sample, MRI, clinical and neuropsychological assessments within 30 days of each other. The exclusion criteria were: 1) having clinical relapse or receiving intravenous corticosteroid therapy within 30 days before the MRI and serum sampling, 2) not able to undergo any of the aforementioned study procedures, and 3) pregnant or nursing mothers.

The CEG-MS study and the retrospective proteomic analyses were approved by the University at Buffalo Institutional Review Board (IRB) and all subjects provided a signed consent form.

### Physical and cognitive disability measures

A board-certified neurologist evaluated patients for global disability using the Expanded Disability Status Scale (EDSS) score^13^, a board certified neuropsychologist oversaw a clinical assessment that included assessment of quantitative mobility and leg function, using the Timed 25 Foot Walk Test (T25FWT)^14^, quantitative finger dexterity using the 9-Hole Peg Test (9HPT)^15^, and cognitive efficiency and speed performance using the Symbol Digit Modalities Test (SDMT) and the Paced Auditory Serial Addition Test (PASAT)^16^. A structured questionnaire was also used to collect demographic and clinical information. According to the clinical presentation and disease history, pwMS were categorized as CIS, relapsing-remitting MS (RRMS), progressive MS (PMS) (further categorized to primary (PPMS), or secondary progressive (SPMS)).^17^

Presence of disability progression (DP) over the follow-up was defined using standard criteria of changes in EDSS scores: 1) An increase of 2 or more points if the baseline EDSS is zero; 2) an increase of 1.5 or more points if the baseline EDSS is 0.5, 3) an increase of ≥1 point if the baseline EDSS is between 1.0 and 5.0, and 4) an increase of equal or greater than 0.5 point if the baseline EDSS is ≥5.5.^18^ Worsening in T25FWT and 9HPT was defined as an increase of greater than or equal to 20% from baseline to follow-up.^14, 15^ Worsening in SDMT performance was defined using several previously used criteria: 1) decrease of 4 or more points from baseline to follow-up ^16^; 2) decrease of 8 or more points from baseline to follow-up; 3) being classified as cognitively impaired based with |z-scores| > 1.5 derived from a healthy control population published in the literature with mean and standard deviation of 55.49, 13.06 respectively ^5, 19^.

### Proteomics analyses

All blood samples were processed within 24-hours from acquisition and stored at −80°C until analyzed. No freeze and thaw cycles were performed in the interim period. All samples were sent to Octave Bioscience (Menlo Park, CA, USA) for proteomic analysis using the MSDA assay panel.^9^ Proteomic analysis was performed blinded to the demographic, clinical and MRI data. The MSDA assay uses Proximity Extension Assay (PEA) methodology and is performed on the Olink^TM^ platform. Twenty-one proteins that are associated with key biological pathways of MS pathophysiology were selected for inclusion on the panel based on results from discovery analyses investigating relative expression of 1196 proteins in previously characterized MS cohorts. The MSDA assay utilizes a stacked classifier logistic regression model of 18 age- and sex adjusted protein concentrations as previously described to determine four disease pathway scores (immunomodulation, neuroinflammation, myelin biology, and neuroaxonal integrity) as well as an overall disease activity score.^9^ The complete list of proteins (with commonly used aliases and their abbreviations) are shown in Figure 1.

**Figure 1.**
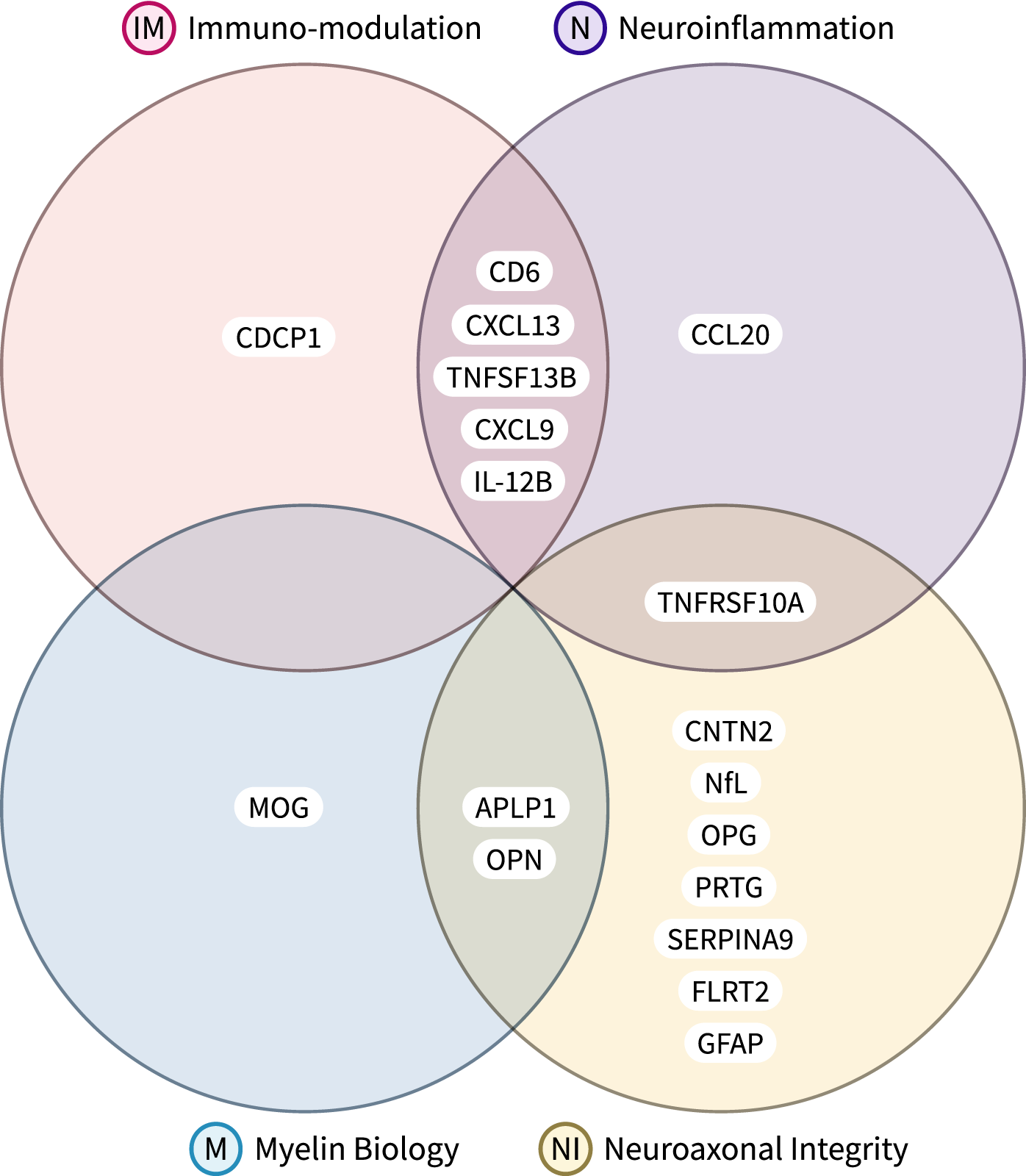
The Octave Bioscience Multiple Sclerosis Disease Activity (MSDA) test was developed using Proximity Extension Assay (PEA) methodology on the Olink™. It measures the concentrations of 18 proteins, and utilizes an algorithm to determine 4 disease pathways scores (immunomodulation, neuroinflammation, myelin biology, and neuroaxonal integrity) and an overall disease activity score.

### MRI acquisition and analyses

At baseline and follow-up visits, pwMS underwent an MRI examination using the same 3T Signa Excite 12 Twin-Speed scanner (GE Healthcare, Milwaukee, WI, USA) and eight channel head and neck coil. The standard sequences utilized in these analyses were two-dimensional (2D) fluid attenuated inversion recovery (FLAIR), 2D T1-weighted spin echo with and without use of 0.2mL/kg gadolinium (Gd) contrast acquired 5 min post-injection, and high-resolution 3D T1-weighted imaging. The sequence parameters are explained in details elsewhere. ^20^

Lesion analysis was performed in a blinded manner with respect to the patient clinical and proteomics status. T2 lesion volume (LV), T1-LV and Gd-LV were obtained using a semi-automated contouring/thresholding technique using Java Image Manipulation (JIM) version 6.0 (Xinapse Systems Ltd, http://www.xinapse.com/, Essex, UK). The cross-sectional and longitudinal changes in volumes of brain regions of interest (ROIs) of whole brain (WB), white matter (WM), gray matter (GM), all normalized for head size, were measured using the SIENAX and SIENA algorithms (FMRIB Software Library, http://www.fmrib.ox.ac.uk/fsl).21 A lesion inpainting technique was used to avoid tissue misclassification.^22^ Total deep GM (DGM) volume and specific volume of the thalamus were obtained with FMRIB’s Integrated Registration and Segmentation Tool (FIRST, https://fsl.fmrib.ox.ac.uk/fsl, version 2.6). The number of patients who had baseline and follow-up samples are described in Supplementary Table 1. Lastly, pathological change in whole brain volume was determined as an annualized percent brain volume reduction of greater than or equal to 0.4%^23^ and pathological lateral ventricle volume change if an annualized percent volume expansion of greater than or equal to 3.5%.^24^

### Statistical analyses

Data and statistical analyses were performed using Python version 3.8.10, SciPy 1.9.3, pandas 1.5.2, pingouin 0.5.3, and NumPy 1.24.1. Ordinary and mixed-effects models were estimated using R version 4.1.3. Logistic regression models were estimated with Python statsmodels package version 0.13.2.

Student’s t-test and Analysis of Covariance (ANCOVA) were used for statistical analysis of parametric continuous variables and longitudinal analysis was performed using the paired non-parametric Wilcoxon test. Ordinary least-squares were used to estimate cross-sectional univariable models; linear mixed-effects regression models were used to estimate models on longitudinal data; linear logistic regression models were used to estimate dichotomous outcomes (e.g., pwMS disability progression yes/no).

MRI-based brain volumes, EDSS, and neuropsychological test outcomes were used as dependent variables, and age, sex, BMI, and all proteomic measures as independent predictors (*outcome score = age + sex + body mass index (BMI) + biomarker concentration*) and for the linear mixed-effects model, subject ID was set as random effect (*outcome score = age + sex + BMI + biomarker concentration + (1|patient ID*). For entry into the regression models, the proteomic data, MRI-based brain volumes, EDSS and neuropsychological test scores were transformed using log(10) and all the statistical tests were applied to the log-transformed data. Logistic regression models were similarly used if the dependent variable was of categorical nature. Outcomes such as R^2^ for ordinary and mixed-effects regression, McFadden’s pseudo-R^2^ for logistic regression, standardized β and p-values were reported. Adjusted p-values lower than 0.05 were considered statistically significant. The regression and correlation p-values underwent false discovery rate (FDR) correction using the Benjamini-Hochberg procedure.

Data were visualized using Python matplotlib 3.4.2, seaborn 0.12.1, and plotly 5.12.0 packages. The data distribution was determined using visual inspection of histograms and Q-Q plots. Volcano plots were used to visualize the significance (p-value) versus effect size.

## Results

### Demographic and clinical characteristics

Table 1 describes the demographics and clinical characteristics of the pwMS. As expected, the pwPMS were significantly older, had longer disease duration and higher EDSS scores both at baseline and follow-up visits (p<0.001 for all). We failed to reject the null hypothesis that the rate of DP between pwCIS/RRMS and pwPMS were similar (28.6% vs. 37.5%, p=0.251). There were no significant differences in terms of baseline and follow-up DMT use. As expected, the pwPMS had significantly greater pathology measured by conventional MRI measures of T2-LV and T1-LV (p<0.001 and p=0.015) and global measures of WBV, WMV and GMV (p<0.001). The pwCIS/RRMS had on average significantly more Gd lesions when compared to the PMS group (p<0.001). Supplement Figure 1 depicts the distribution of disease phenotypes at baseline and transition over the follow-up.

**Table 1.** Demographic, clinical and conventional MRI characteristics of the study population.

| Demographic and clinical characteristics | pwMS<br>(n=202) | CIS/RRMS<br>(n=148) | PMS<br>(n=54) | p-value |
| --- | --- | --- | --- | --- |
| Female, n (%) | 151 (74.8) | 106 (71.6) | 45 (83.3) | 0.09<sup>a</sup> |
| Age at baseline, mean (SD) | 47.1 (11.1) | 44.1 (10.6) | 55.3 (7.9) | <0.001<sup>b</sup> |
| Time of follow-up, mean (SD) | 5.4 (0.6) | 5.4 (0.6) | 5.5 (0.6) | 0.732<sup>b</sup> |
| BMI at baseline, mean (SD) | 27.5 (5.8) | 27.9 (6.2) | 26.5 (4.5) | 0.1<sup>b</sup> |
| Age of disease onset, mean (SD) | 32.9 (9.8) | 32.6 (9.0) | 33.6 (11.8) | 0.6<sup>b</sup> |
| Disease duration at baseline, mean (SD) | 13.4 (10.2) | 11.1 (8.5) | 21.7 (10.5) | <0.001<sup>b</sup> |
| EDSS at baseline, median (IQR) | 2.5 (1.5-5.0) | 1.5 (1.5-2.5) | 6.0 (4.0-6.5) | <0.001<sup>c</sup> |
| EDSS at follow-up, median (IQR) | 3.0 (1.6-6.0) | 2.0 (1.5-3.5) | 6.5 (4.0-6.5) | <0.001<sup>c</sup> |
| EDSS absolute change, mean (SD) | 0.4 (0.9) | 0.4 (0.9) | 0.4 (0.7) | <0.001<sup>b</sup> |
| Disability progression, n (%)* | 56 (30.9) | 38 (28.6) | 18 (37.5) | 0.251<sup>a</sup> |
| Relapse rate over the follow-up, mean (SD) | 0.172 (0.369) | 0.204 (0.4) | 0.09 (0.24) | <0.001<sup>d</sup> |
| DMT at baseline, n (%) |  |  |  |  |
| IFN-β | 85 (42.1) | 60 (40.5) | 25 (46.3) | 0.271<sup>a</sup> |
| Glatiramer acetate | 37 (18.3) | 24 (16.2) | 13 (24.1) |  |
| Natalizumab | 29 (14.4) | 25 (16.9) | 4 (7.4) |  |
| Off-label DMT | 5 (2.5) | 3 (2.0) | 2 (3.7) |  |
| No DMT | 46 (22.8) | 36 (24.3) | 10 (18.5) |  |
| DMT at follow-up, n (%) |  |  |  |  |
| IFN-β | 68 (33.7) | 52 (35.1) | 16 (29.6) | 0.797<sup>a</sup> |
| Glatiramer acetate | 45 (22.3) | 31 (20.9) | 14 (25.9) |  |
| Natalizumab | 15 (7.4) | 12 (8.1) | 3 (5.6) |  |
| Oral DMT | 28 (13.9) | 22 (14.9) | 6 (11.1) |  |
| Off-label DMT | 12 (5.9) | 8 (5.4) | 4 (7.4) |  |
| No DMT | 34 (16.8) | 23 (15.5) | 11 (20.4) |  |
| T2-LV, mean (SD) | 13.5 (16.7) | 10.3 (14.09) | 22.2 (20.1) | <0.001<sup>b</sup> |
| T1-LV, mean (SD) | 3.0 (7.2) | 2.2 (6.44) | 5.5 (8.6) | 0.015<sup>b</sup> |
| Gd-LN, mean (SD) | 0.05 (3.2) | 0.7 (3.7) | 0.04 (0.2) | <0.001<sup>c</sup> |
| Gd-LV, mean (SD) | 0.07 (0.4) | 0.1 (0.48) | 0.01 (0.03) | 0.194<sup>b</sup> |
| WBV, mean (SD) | 1466.4 (94.3) | 1490.2 (87.7) | 1401.7 (80.8) | <0.001<sup>b</sup> |
| WMV, mean (SD) | 725.9 (62.2) | 738.7 (62.1) | 689.8 (46.7) | <0.001<sup>b</sup> |
| GMV, mean (SD) | 740.8 (63.9) | 751.4 (65.9) | 711.9 (48.1) | <0.001<sup>b</sup> |

**Legend:** MS – multiple sclerosis, CIS – clinically isolated syndrome, RRMS – relapsing remitting multiple sclerosis, PMS – progressive multiple sclerosis, BMI – body mass index, EDSS – Expanded Disability Status Scale, DMT – disease modifying therapy, IFN – interferon, SD – standard deviation, IQR – interquartile range. LV – lesion volume, LN – lesion number, WBV – whole brain volume, WMV – white matter volume, GMV – gray matter volume. Thirteen (13) CIS/RRMS patients transitioned into PMS over the follow-up. Parametric data is shown as mean (standard deviation), whereas non-parametric data is shown as median (interquartile range). The specific comparisons were performed using; a – $\chi^2$ test, b – Student's t-test, c – Mann Whitney U test, d – Negative binomial regression. P-values lower than 0.05 were considered statistically significant and shown in bold. <sup>\*</sup>- Disability progression was available for 181 out of 202 pwMS due to missing EDSS values at either baseline or follow-up visit.

Out of the pwMS with available longitudinal disability data, 55 out of 181 (30.4%) worsened in EDSS scores, 46 out of 186 (24.7%) had a pathological rate of whole brain atrophy and only 19 out of 186 (10.2%) has a pathological rate of ventricle enlargement. The paired statistical test revealed that all outcomes worsened significantly at the follow-up time-point except for the WMV and PASAT performance (Supplement Table 2). Supplement Figure 2 visualizes shifts in MRI-based volumes, EDSS and neuropsychological scores between the baseline and follow-up time-points. Supplement Figure 3, 4 demonstrate differences in MRI-based volumes, EDSS, neuropsychological scores and biomarker concentrations between pwCIS/RRMS and pwPMS subgroups.

### Proteomic characteristics of the study population

In total, 202 pwMS had serum samples at the baseline visit and 143 pwMS had serum samples at both the baseline and follow-up visits. The baseline, follow-up and longitudinal change in each of the proteomic biomarkers (shown as median and interquartile range (IQR)) are shown in Figure 2 for biomarkers with significant shift between the time points and detailed analysis is shown in Supplementary Table 4. Over the follow-up, pwMS had a significant 56% increase in CCL20 (9.74pg/mL vs. 15.1pg/mL, p=0.001) and 9.4% increase in NfL (10.5 pg/mL vs 11.5 pg/mL, p=0.003). There were also significant shifts in CDCP1, CNTN2, GFAP, MOG, and OPN but they did not survive multiple comparisons correction. Supplement Figure 4 demonstrates significant shifts in the serum biomarkers between pwCIS/RRMS and pwPMS at baseline and follow-up time-points.

**Figure 2.**
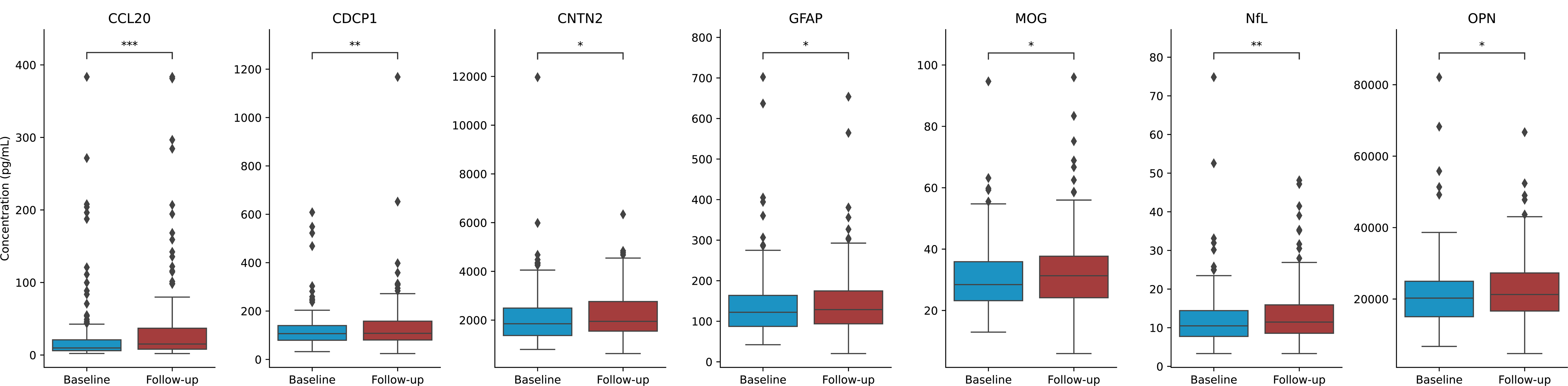
Changes in blood serum biomarker concentration between the baseline and follow-up time points for those with p-value < 0.05. **Legend:** Paired Wilcoxon signed-rank test was used to compare between baseline and follow-up time points. P-value annotation legend: ns: 5.00e-02 < p <= 1.00e+00, *: 1.00e-02 < p <= 5.00e-02, **: 1.00e-03 < p <= 1.00e-02, ***: 1.00e-04 < p <= 1.00e-03, ****: p <= 1.00e-04.

In the pwMS who progressed over the follow-up, there were several proteins whose levels differed between the visits. For the patients who progressed in the 9HPT test, CDCP1 increased by 31% (110 pg/ml vs. 144 pg/ml, p=0.003), TNFRSf10A increased by 37% (5.61 pg/ml vs 7.71 pg/ml, p=0.005) and VCAN increased by 17% (428 pg/ml vs 500 pg/ml, p=0.007). The other proteins in Table 2 did not survive the multiple comparisons correction. Although age adjusted NfL was increased and PRTG was decreased in patients with progressive type at follow-up, these findings did not pass the multiple comparisons correction.

**Table 2.** Changes in blood serum protein concentrations between baseline and the follow-up for pwMS with worsening in disability measures.

| Endpoint | Biomarker | Baseline median (IQR) | Follow-up median (IQR) | Percentage Change (%) | p-value |
| --- | --- | --- | --- | --- | --- |
| EDSS worsening | CCL20 | 11.5 (6.12, 19.3) | 15.9 (10.3, 41.5) | 38 | **0.041** |
|  | CDCP1 | 108 (81.9, 126) | 135 (86.7, 180) | 25 | **0.005** |
|  | TNFSF13B | 4.82 (4.0, 5.95) | 5.19 (4.51, 6.27) | 7.6 | **0.008** |
| 20% 9HPT worsening | CCL20 | 12.4 (7.68, 19.1) | 28.1 (13.9, 42.0) | 130 | **0.027** |
|  | CDCP1 | 110 (105, 125) | 144 (112, 193) | 31 | **0.003\*** |
|  | CXCL13 | 51.7 (37.8, 78.1) | 61.9 (51.0, 103) | 20 | **0.012** |
|  | CXCL9 | 55.7 (39.1, 77.8) | 69.1 (43.9, 101) | 24 | **0.042** |
|  | OPN | 20.9 (16.6, 30.6) | 25.2 (19.4, 33.5) | 20 | **0.042** |
|  | TNFRSF10A | 5.61 (4.66, 7.65) | 7.71 (6.51, 8.5) | 37 | **0.005\*** |
|  | VCAN | 428 (393, 473) | 500 (460, 571) | 17 | **0.007\*** |
| 4 Points SDMT Worsening | MOG | 29.9 (23.0, 38.5) | 32.0 (25.2, 43.4) | 6.9 | **0.027** |
| 8 Points SDMT Worsening | SERPINA9 | 61.3 (34.3, 77.4) | 32.1 (16.1, 59.3) | -48 | **0.01** |
| %0.4 loss in WBV | CDCP1 | 105 (66.3, 132) | 107 (83.7, 147) | 1.5 | **0.014** |
| 3.5% increase in LV | SERPINA9 | 73.0 (57.1, 89.5) | 64.4 (38.2, 76.6) | -12 | **0.049** |

**Legend:** EDSS – Expanded Disability Status Scale, 9HPT – 9-Hole Peg Test, WBV – whole brain volume.
Wilcoxon signed-rank tested the significance in shifts. P-values smaller than 0.05 were considered significant and are highlighted with bold fonts, and those with asterisks (\*) survived the Benjamini-Hochberg correction for false discovery rate (FDR). All measures are shown as pg/mL except for TNFSF13B and OPN that are shown as ng/mL.

### Relationship between proteomic data and outcomes in pwMS

#### Longitudinal models

Figure 3 shows the significant biomarkers of the mixed-effects models estimated for both time-points and all pwMS. We found six biomarker candidates, namely GFAP, FLRT2, CDCP1, TNFRSF10A, CXCL9 and CCL20, whose increases were correlated with reduction in brain volumes and one protein (APLP1) whose decrease was correlated with reduction in brain volume. In particular, GFAP (p=0.003), FLRT2 (p=0.001), CDCP1 (p=0.004) and TNFRSF10A (p=0.003) were associated with WBV decrease and survived the multiple comparisons correction. For the WMV, GFAP was the only protein that survived the statistical test (p<0.001). For the DGM, significant associations with CCL20 (p=0.004), CXCL9 (p=0.001), CDCP1 (p=0.002), FLRT2 (p=0.008), TNFRSF10A (p=0.009) were noted.

**Figure 3.**
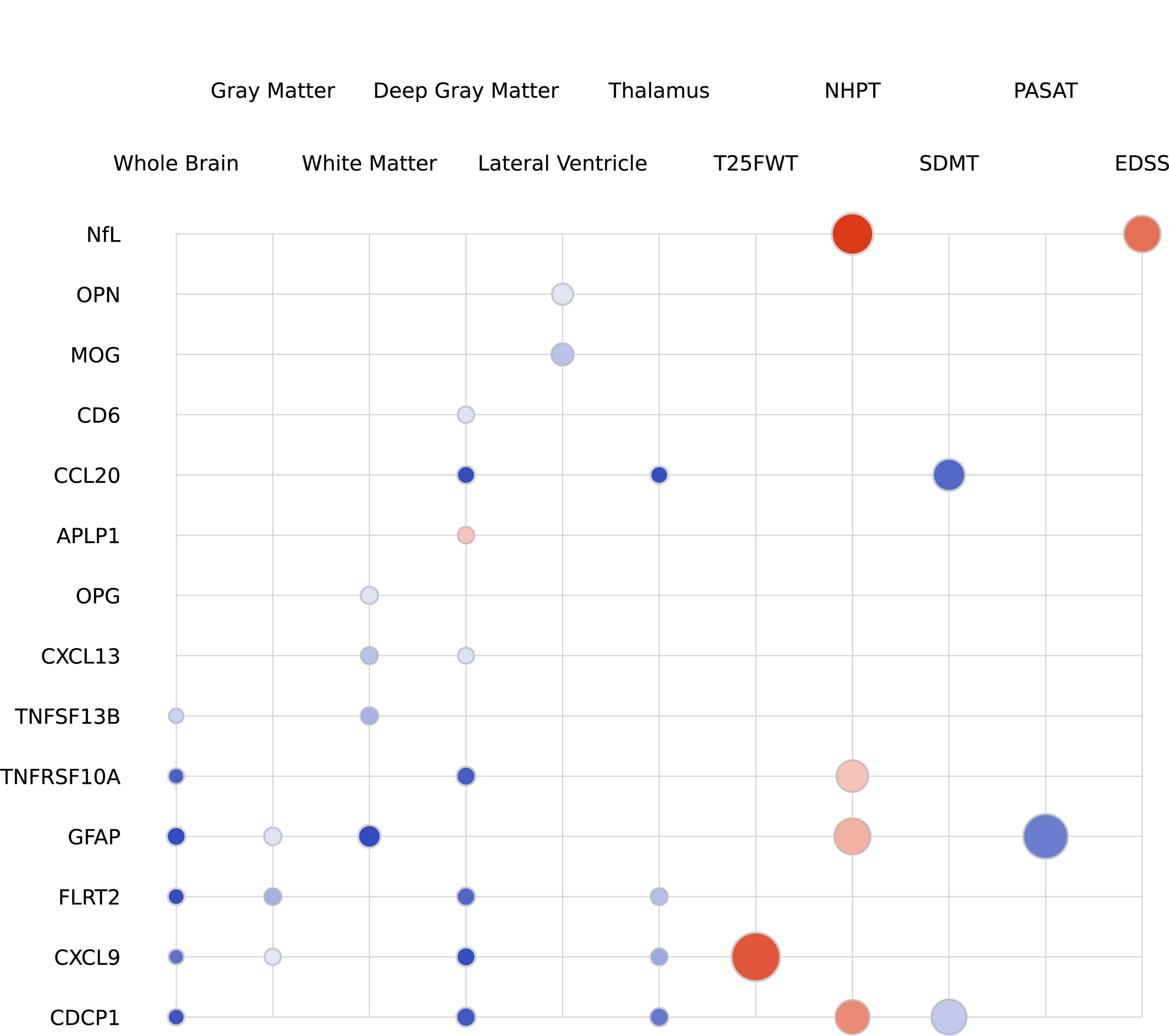
Longitudinal single-protein model parameters with adjustment for age, sex and BMI. **Legend:** The radius of each circle is proportional to the estimated standardized coefficient of the corresponding protein, red (blue) circles represent proteins with positive (negative) effects in estimating the second-class label. The opacity of each circle represents the p-value; a p-value of < 0.001 corresponds to full opacity and a p-value of 0.05 corresponds to the least opacity. Biomarkers that survived the multiple comparisons correction are marked with a gold star (*).

In terms of clinical (EDSS scores) and cognitive assessment, we found that worsening in the 9-HPT score was associated with an increase in NfL (p=0.001) surviving the multiple comparisons correction. Table 3 details the estimated coefficients, p-values and the qualities of the fit (R-squared of the model with standardized features as biomarker, sex, age, BMI) corresponding to each biomarker and outcome measure. Note that only significant biomarkers (p<0.05) are included in Table 3.

**Table 3.** Parameters of the linear mixed-effects model predicting outcome score using single-protein models consisting of a biomarker protein concentration, age, sex, BMI and time-point that passed the significant threshold of p<0.05.

| Endpoint | Biomarker | Estimate | R-squared | P-value |
| --- | --- | --- | --- | --- |
| **DGMV** | CD6 | -0.003 | 0.15 | 0.035 |
|  | TNFRSF10A | -0.004 | 0.17 | 0.009* |
|  | MOG | 0.003 | 0.18 | 0.045 |
|  | FLRT2 | -0.004 | 0.19 | 0.008* |
|  | CXCL9 | -0.005 | 0.16 | 0.001* |
|  | CDCP1 | -0.005 | 0.18 | 0.002* |
|  | CCL20 | -0.003 | 0.16 | 0.004* |
|  | APLP1 | 0.004 | 0.17 | 0.011* |
| **GMV** | FLRT2 | -0.004 | 0.27 | 0.017 |
|  | CXCL9 | -0.003 | 0.25 | 0.04 |
|  | CCL20 | -0.003 | 0.26 | 0.006 |
| **LVV** | MOG | -0.008 | 0.14 | 0.031 |
|  | OPN | -0.007 | 0.14 | 0.041 |
| **Thalamus** | CCL20 | -0.003 | 0.16 | 0.006 |
|  | CXCL9 | -0.003 | 0.15 | 0.031 |
|  | FLRT2 | -0.003 | 0.18 | 0.037 |
|  | CDCP1 | -0.005 | 0.17 | 0.007 |
| **WMV** | GFAP | -0.008 | 0.12 | <0.001* |
|  | TNFSF13B | -0.003 | 0.11 | 0.046 |
| **WBV** | CDCP1 | -0.003 | 0.24 | 0.004* |
|  | CXCL9 | -0.002 | 0.23 | 0.014 |
|  | FLRT2 | -0.003 | 0.25 | 0.001* |
|  | GFAP | -0.004 | 0.28 | <0.001* |
|  | OPG | -0.002 | 0.23 | 0.05 |
|  | TNFSF13B | -0.002 | 0.24 | 0.028 |
|  | TNFRSF10A | -0.003 | 0.24 | 0.003* |
| **EDSS** | NfL | 0.023 | 0.24 | 0.021 |
| **9HPT** | NfL | 0.031 | 0.21 | 0.001* |
|  | GFAP | 0.022 | 0.2 | 0.037 |
|  | CDCP1 | 0.02 | 0.16 | 0.028 |
| **PASAT** | GFAP | -0.038 | 0.07 | 0.012 |
|  | CCL20 | -0.018 | 0.02 | 0.042 |
| **SDMT** | CDCP1 | -0.021 | 0.11 | 0.042 |
|  | CCL20 | -0.017 | 0.09 | 0.008 |
| **T25FWT** | CXCL9 | 0.041 | 0.08 | 0.014 |

**Legend:** pwMS – people with multiple sclerosis, DGMV – deep gray matter volume, GMV – gray matter volume, LVV – lateral ventricular volume, WMV – white matter volume, WBV – whole brain volume, EDSS – Expanded Disability Status Scale, 9HPT – 9-Hole Peg Test, PASAT – Paced Auditory Serial Addition Test, SDMT – Symbol Digit Modalities Test, T25FWT – Timed 25-Foot Walk Test. P-values smaller than 0.05 were considered significant and are highlighted with bold fonts, and those with asterisks (\*) survived the Benjamini-Hochberg correction for false discovery rate (FDR).

#### Cross-sectional models

Figure 4 shows the significant biomarkers of linear single-protein models estimated individually for the baseline and follow-up outcomes. At the baseline visit, higher GFAP was correlated with lower WBV (p<0.001), GMV (p<0.001), thalamic volume (p<0.001), and DGMV (p<0.001), and higher LVV (p<0.001). Moreover, higher GFAP was associated with higher EDSS scores (p=0.002). Detailed analyses are shown in Table 4.

**Figure 4.**
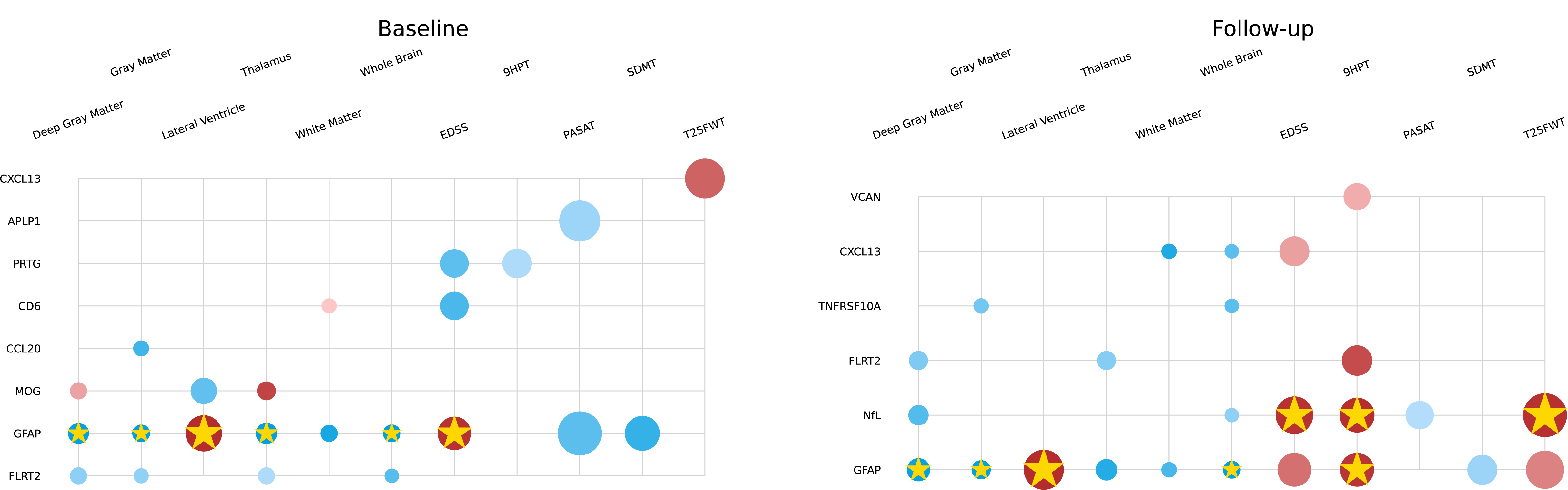
Cross-sectional single-protein model parameters with adjustment for age, sex and BMI for baseline (left) and follow-up (right). **Legend:** The radius of each circle is proportional to the estimated standardized coefficient of the corresponding protein, red (blue) circles represent proteins with positive (negative) effects in estimating the second-class label. The opacity of each circle represents the p-value; a p-value < 0.001 corresponds to full opacity and a p-value of 0.05 corresponds to the least opacity. Biomarkers that survived the multiple comparisons correction are marked with a gold star (*).

**Table 4.**
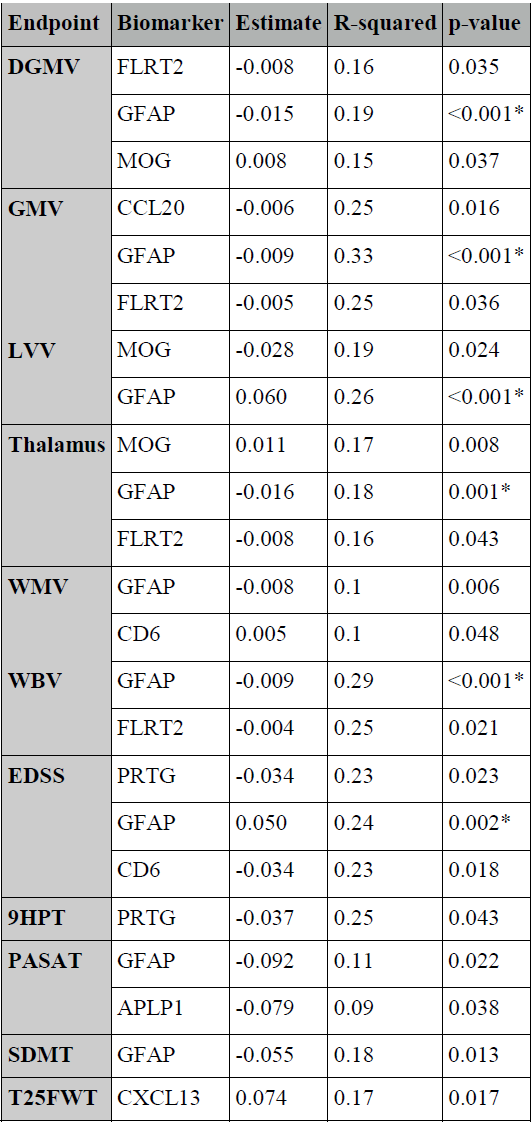
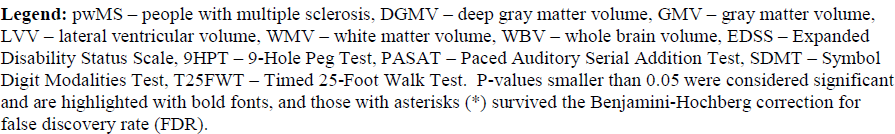
Parameters of the linear model predicting baseline outcome score using single-protein models consisting of <u>baseline</u> biomarker protein concentration, age, sex, BMI that passed the p=0.05 significance threshold.

At the follow-up GFAP, was correlated with lower WBV (p=0.001), GMV (p=0.001), and DGMV (p=0.002), and higher LVV (p<0.001). Both NfL (p=0.001) and GFAP (p=0.003) were correlated with worse 9HPT scores. NfL was also correlated with T25FWT (p=0.002) and EDSS scores (p=0.002). Detailed analyses are shown in Supplement Table 5.

#### Shifts in biomarker levels as predictors of clinical change

It is of importance to assess whether shifts in the biomarker concentration is correlated with shifts in outcome scores. Therefore, we fitted a model with shifts in biomarker between baseline and follow-up, age, sex, BMI as independent predictors and shifts in outcome scores as dependent variables. CDCP1 (p=0.001), FLRT2 (p=0.001), PRTG (p=0.008), TNFRSF10A (p<0.001), TNFSF13B (p<0.001) were predicted shift in the WBV. CD6 (p=0.001) was predicted shift in the DGMV. Figure 5 and Supplement Table 6 describe the findings of these analyses.

**Figure 5.**
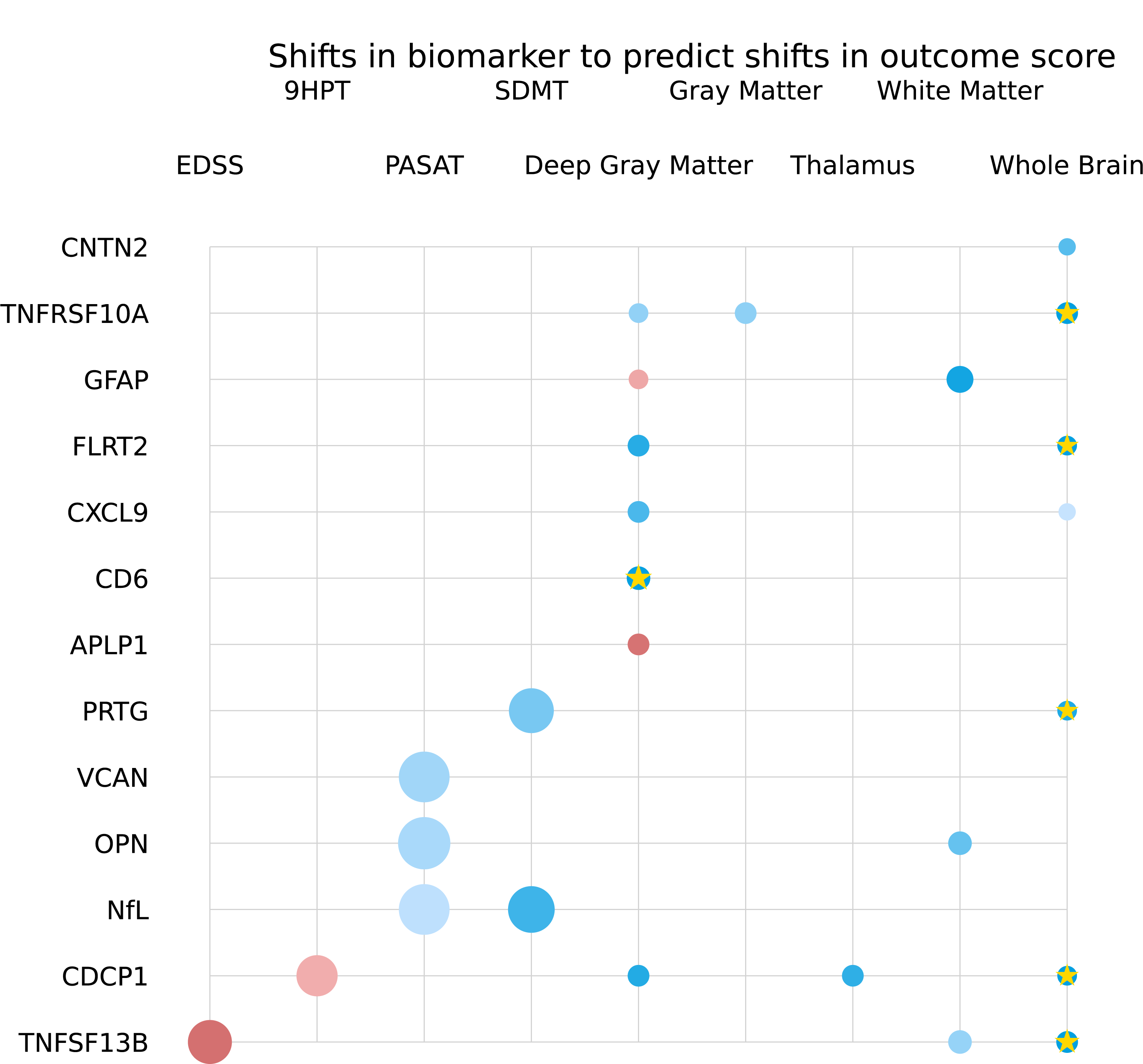
Single-protein model parameters predicting shifts in outcome score using shifts in biomarker concentration with adjustment for age, sex and BMI. **Legend:** The radius of each circle is proportional to the estimated standardized coefficient of the corresponding protein, red (blue) circles represent proteins with positive (negative) effects in estimating the second-class label. The opacity of each circle represents the p-value; a p-value < 0.001 corresponds to full opacity and a p-value of 0.05 corresponds to the least opacity. Biomarkers that survived the multiple comparisons correction are marked with a gold star (*).

## Discussion

The findings of this longitudinal proteomics study are multifold. Firstly, multiple proteomic biomarkers representing different pathophysiological MS pathways are differentially associated with phenotypical and macroscopic pathological changes. Secondly, worse physical and cognitive outcomes in pwMS were associated with blood-based measures of NfL. Lastly, worse neurodegenerative MRI outcomes were associated with a greater number of biomarkers including GFAP, CDCP1, CXCL9, CCL20, APLP1, FLRT2, аnd TNFRSF10A.

The NfL/GFAP relationship with clinical outcomes was recently demonstrated in a similar longitudinal Swiss study.^25^ Over an average follow-up of 7 years, serum GFAP levels were prognostic of progression independent of relapse activity (PIRA) and complementary to the serum NfL data.^25^ Moreover, NfL levels were prognostic of atrophy in the WMV, whereas GFAP specifically prognosticate GM atrophy.^25^ The multi-protein panel employed in our study was also utilized in a study of 431 unique pwMS and successfully predicted the real-world disability status (patient-reported disability score and patient-reported outcomes).^26^ The proteomic profiles consistently outperformed individual top-ranking markers such as NfL and GFAP.^26^ The fact that the same protein biomarkers implicated in their stacking classification algorithm (CDCP1, IL-12B and PRTG) were also seen in our objective disability findings further validates the utility and need of multi-protein proteomic assay.^26^

We further expand on the literature by demonstrating that the similar set of biomarkers (APLP1, CDCP1, FLRT2, TNFRSF10A and CCL20) are also relevant to MRI-based volumetric measures. Of note, the directionality of APLP1-DGMV relationship (decrease in the biomarker concentration was associated with decreased DGMV) was opposite when compared to the remaining ones. Currently, there are no comprehensive proteomic studies that investigate associations with MRI measures in pwMS. The literature most commonly describes individual associations with one or two proteomic measures (NfL and GFAP).^20, 27, 28^ Despite the high collinearity between serum NfL and GFAP levels, a cross-sectional study of 129 pwMS showed that the amount of lesion pathology (T2-LV) and WM/GM volumes were associated only with GFAP levels and not with NfL.^29^ We corroborate these findings with GFAP remaining a strong predictor all MRI measures acquired in our study (WBV, DGM, LVV and thalamic volume). Serum GFAP levels were also recently associated with greater microstructural pathology in 62 pwMS assessed by diffusion tensor imaging.^30^ Our results showed that CCL20 has strong association with the DGM volume and it was increased at the follow-up time point. This protein was previously shown to be increased in PwMS and specifically with a progression index and was higher during remission than in relapse periods. ^31, 32^

The multiplex assays could broaden our understanding of key mechanisms underlying progression by taking a biological-based approach to objectively quantify disease progression.^33^ They have been demonstrated in other neurological disorders as well.^34, 35^ For example, proteomic data from only 4-Plex assay (NfL, GFAP, tau protein and ubiquitin c-terminal hydrolase L1; UCH-L1) better classified people with traumatic brain injury when compared to only single proteomic measure.^34^ Similarly, cognitive performance in Alzheimer’s Disease and amyloid PET status can be predicted and classified by a combination of GFAP, amyloid beta and neurofilament light chain.^35^

While cut-offs of normal versus pathological levels of NfL in pwMS have been previously published ^36, 37^, this information is not available for the majority of proteomic biomarkers utilized in this multi-protein assay. A limited number of studies report the reference intervals and preanalytical GFAP levels.^38, 39^ For example, a Danish-based analysis of 371 apparently healthy subjects reported fairly large ranges with GFAP levels of 25-136 ng/L (20-39 years old), 34-242 ng/L (40-64 years old) and 4-438 ng/L (for 65-90 years old).^38^ Moreover, there was ∼10% variability after three freeze-thaw cycles or storing serum samples at −20 °C for an average of 133 days.^38^ Significant semidiurnal variations in GFAP have been reported (9 AM vs. 12 PM vs 9 PM blood draw).^40^ Based on these references, none of the median pwMS values would be considered “pathological”. Other biomarkers such as contactin-1 may be more susceptible to pre-analytical factors and have even greater variability.^41^ After the selection of best performing biomarkers and creation of multi-protein scores, future studies should aim at determining appropriate cut-offs for best differentiation between normal and pathological states.

The linear modeling approach taken in this study can be considered as a study limitation. Since there is no guarantee that the underlying interaction is inherently linear, an area of future work is to consider nonlinear regression techniques to capture more complex interactions of serum biomarker concentrations and metrics of disease progression. In addition, multi-protein regression approaches in cross-validation studies can be considered to integrate the predictive power of individual biomarkers in one model.^42^ The shifts in DP outcome measures were not uniform. Such imbalance in the training data can deteriorate generalizability of the model.^43, 44^ In the future, it will be of interest to balance the data using up, down techniques or using weighted learners. The significantly lower number of available cognitive measures (only 25% of the pwMS) for the baseline time point presents as another study limitation. Moreover, the initial determination of biomarkers within the assay were based on ability to predict presence of contrast-enhancing and new/newly enlarging lesions.^9^ In comparison to younger more active pwMS from the literature, our population was relatively older and had very limited neuroinflammatory activity. Future development of a more comprehensive assay that contains proteins specific to neurodegenerative changes (vs. neuroinflammation) could better predict the occurrence of long-term disability worsening. Moreover, the use and change in DMT should be incorporated in future statistical analyses. Additional limitation of our analysis is the lack of a third clinical visit that would allow confirmation of the disease progression. Lastly, the lack of healthy control data does not allow us to determine pathological protein cut-offs and risk stratify the pwMS based on healthy condition.

In conclusion, the clinical, cognitive, and MRI-based outcomes in pwMS are associated with more than one proteomic biomarker. While sNfL had the strongest associations with physical disability such as EDSS scores and hand dexterity, additional proteomic biomarkers related to neuroaxonal integrity were associated with cross-sectional and longitudinal MRI measures of brain atrophy. Multi-protein assays may be essential in capturing the complex MS pathophysiology as part of the disease stratification and monitoring. Before implementation into routine clinical practice, future studies should determine the treatment responsiveness of such proteomic biomarkers.

## Data Availability

All data produced in the present work are contained in the manuscript and supplementary materials.

## Acknowledgements

This research has been supported by Octave Bioscience Inc.

## Author Contributions

KJ, AK, SM, FQ, AG contributed to the analysis of data and drafting a significant portion of the manuscript or figures

