## Supplementary Figures for "Proteomic predictors of physical, cognitive and imaging outcomes in multiple sclerosis: 5-year follow-up study"

**Supplement Figure 1.** Patients might change disease course from baseline (left) to follow-up (right) time points.

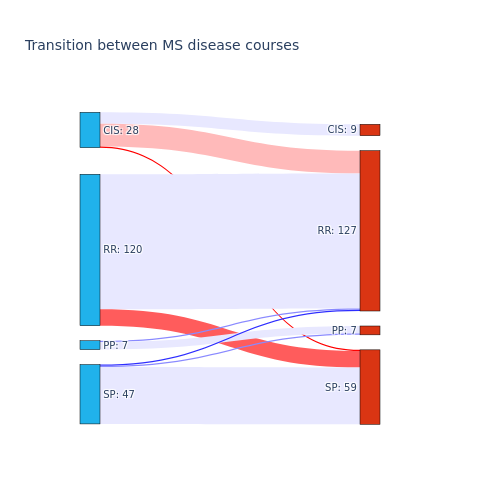

**Legend:** RRMS - relapse remitting multiple sclerosis, CIS - clinically isolated syndrome, SPMS - secondary progressive multiple sclerosis, PPMS - primary progressive multiple sclerosis, RSP - remitting secondary progressive multiple sclerosis.

**Supplement Figure 2.** Changes in brain MRI scan volumes, Expanded Disability Status Scale (EDSS), and neuropsychological scores between baseline and over the follow-up.

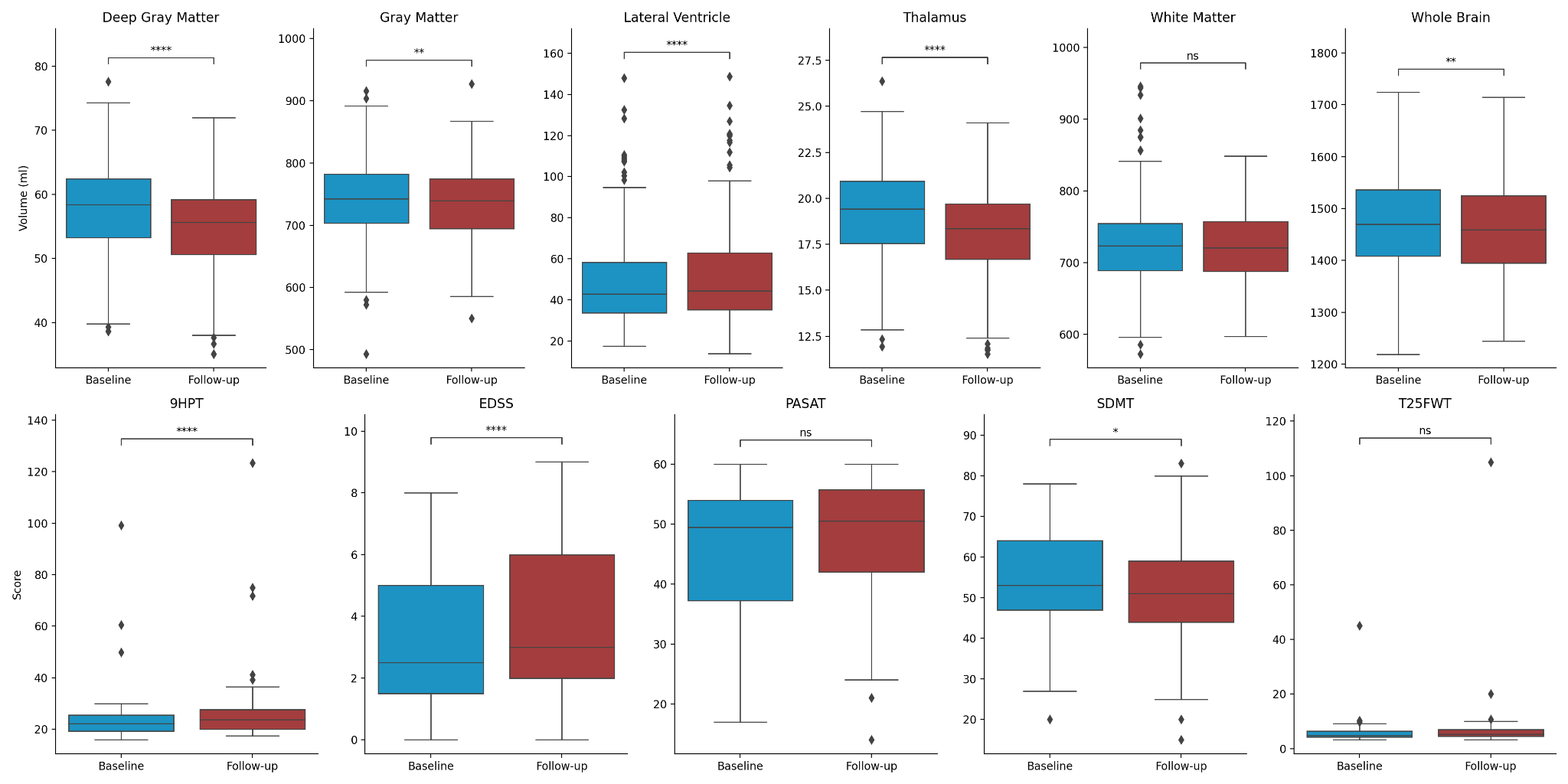

**Legend:** Paired Wilcoxon signed-rank test was used to compare between baseline and follow-up time points. p-value annotation legend: ns: 5.00e-02 < p <= 1.00e+00, *: 1.00e-02 < p <= 5.00e-02, **: 1.00e-03 < p <= 1.00e-02, ***: 1.00e-04 < p <= 1.00e-03, ****: p <= 1.00e-04.

**Supplement Figure 3.** Changes in brain MRI scan volume, Expanded Disability Status Scale (EDSS), and neuropsychological scores between patients with Progressive MS (PMS) and Clinically Isolated Syndrome (CIS) and Relapse Remitting MS(RRMS).

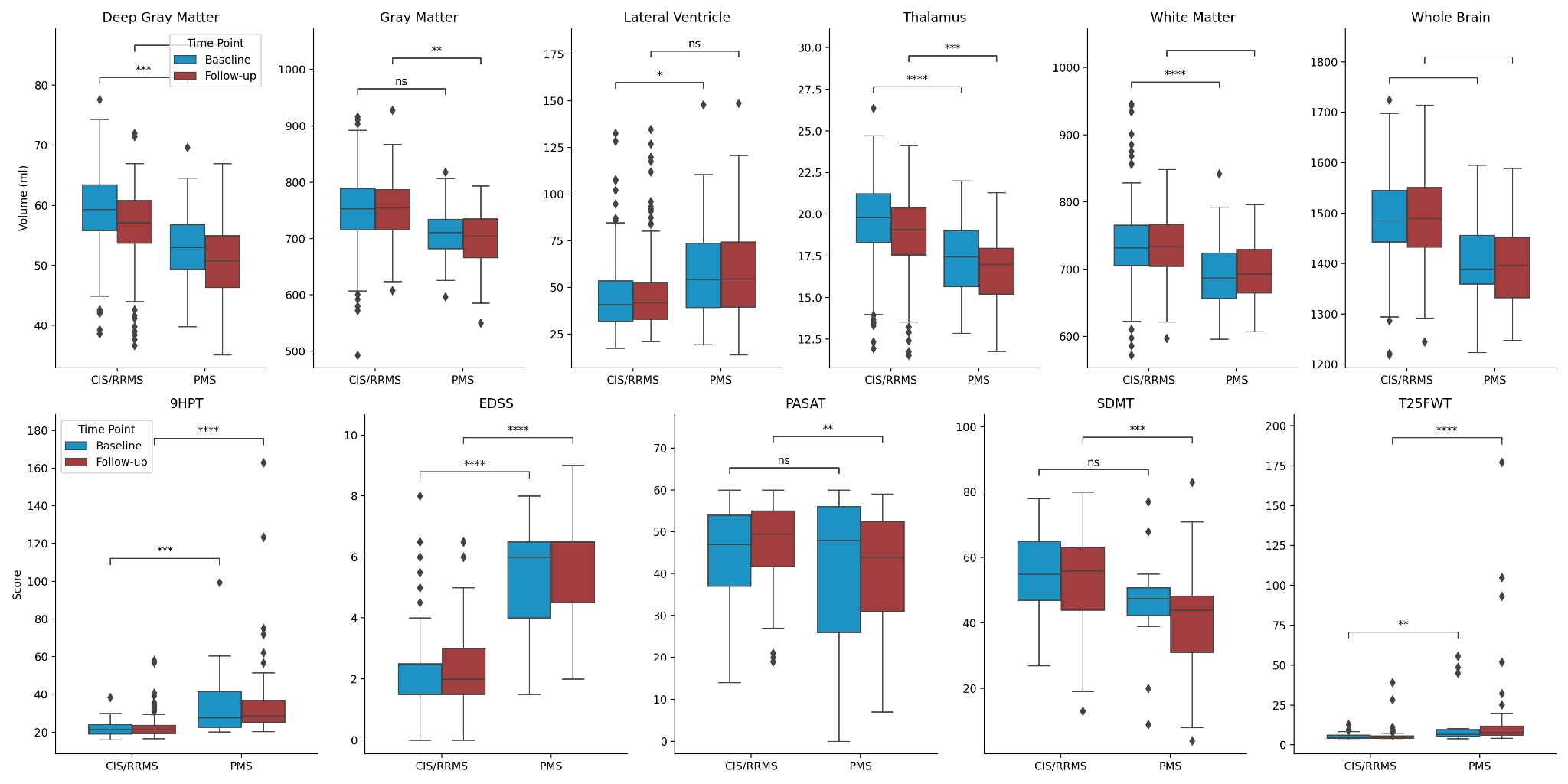

**Legend:** ANalysis of COVAriance (ANCOVA) was used to compare between the two subgroups. p-value annotation legend: ns: 5.00e-02 < p <= 1.00e+00, *: 1.00e-02 < p <= 5.00e-02, **: 1.00e-03 < p <= 1.00e-02, ***: 1.00e-04 < p <= 1.00e-03, ****: p <= 1.00e-04.

**Supplement Figure 4.** Changes in blood serum biomarker concentration between patients with Progressive MS (PMS) and Clinically Isolated Syndrome (CIS) and Relapse Remitting MS (RRMS).

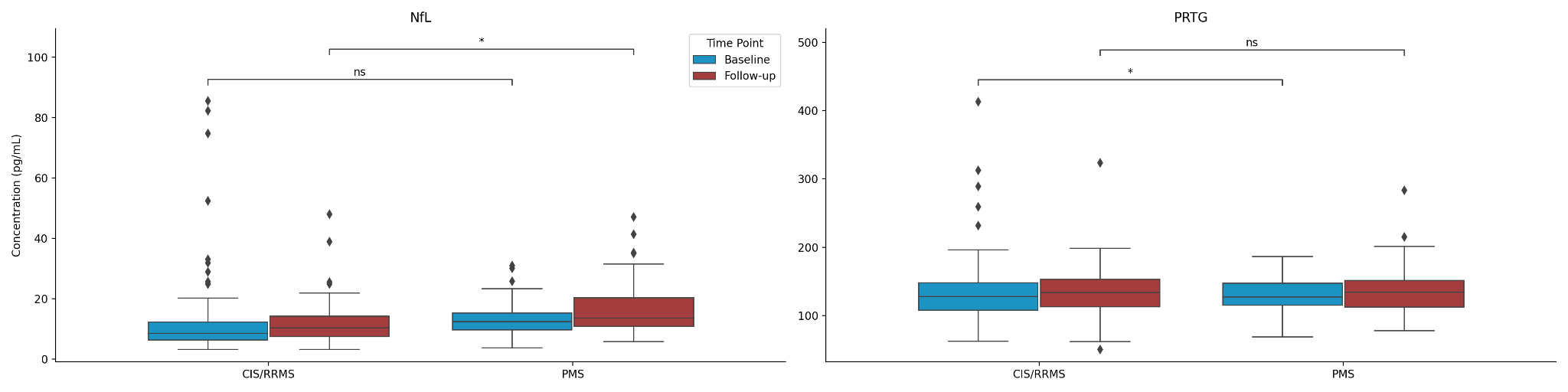

ANalysis of COVAriance (ANCOVA) was used to compare between the two subgroups. p-value annotation legend: ns: 5.00e-02 < p <= 1.00e+00, *: 1.00e-02 < p <= 5.00e-02, **: 1.00e-03 < p <= 1.00e-02, ***: 1.00e-04 < p <= 1.00e-03, ****: p <= 1.00e-04.
