## Supplementary Tables for "Proteomic predictors of physical, cognitive and imaging outcomes in multiple sclerosis: 5-year follow-up study"

**Supplement Table 1.** Number of pwMS with blood samples and demographic information per time-point and endpoint.

| **Endpoint** | **Time Point** | **N** | **Age** | **BMI** | **Female** % |
| --- | --- | --- | --- | --- | --- |
| **MRI measures** | baseline | 201 | 47.1 (11.1) | 27.5 (5.73) | 74.6 |
|  | follow-up | 135 | 53.8 (10.9) | 27.3 (5.64) | 74.1 |
| **EDSS** | baseline | 186 | 46.9 (10.9) | 27.5 (5.63) | 73.7 |
|  | follow-up | 138 | 53.6 (11.1) | 27.5 (5.84) | 73.9 |
| **9HPT** | baseline | 57 | 46.7 (11.0) | 27.3 (5.45) | 71.9 |
|  | follow-up | 134 | 54.0 (10.7) | 27.7 (5.77) | 73.1 |
| **PASAT** | baseline | 58 | 45.9 (11.2) | 27.5 (5.66) | 72.4 |
|  | follow-up | 132 | 54.0 (10.7) | 27.6 (5.84) | 72.0 |
| **SDMT** | baseline | 59 | 46.1 (11.3) | 27.6 (5.61) | 72.9 |
|  | follow-up | 138 | 54.3 (10.7) | 27.6 (5.74) | 73.2 |
| **T25FWT** | baseline | 59 | 45.9 (11.5) | 27.3 (5.42) | 72.9 |
|  | follow-up | 129 | 53.6 (10.9) | 27.5 (5.90) | 72.1 |

**Legend:** pwMS – people with multiple sclerosis, MRI – magnetic resonance imaging, EDSS – Expanded Disability Status Scale, 9HPT – 9-Hole Peg Test, PASAT – Paced Auditory Serial Addition Test, SDMT – Symbol Digit Modalities Test, T25FWT – Timed 25-Foot Walk Test.

### **Supplement Table 2.** Changes in brain MRI scan volume, Expanded Disability Status Scale (EDSS), and neuropsychological scores between baseline and over the follow-up.

| **Endpoint** | **N of patients with longitudinal data** | **Baseline median (IQR)** | **Follow-up median (IQR)** | **Percentage Change (%)** | **P-value** |
| --- | --- | --- | --- | --- | --- |
| DGMV | 185 | 58.4 (53.3, 62.4) | 55.6 (50.6, 59.2) | -4.8 | **<0.001*** |
| GMV | 186 | 742 (703, 782) | 739 (694, 775) | -0.42 | **0.002*** |
| LVV | 186 | 42.8 (33.8, 58.3) | 44.3 (35.2, 62.8) | 3.4 | **<0.001*** |
| Thalamus | 185 | 19.4 (17.5, 20.9) | 18.4 (16.7, 19.7) | -5.4 | **<0.001*** |
| WMV | 186 | 724 (689, 755) | 721 (688, 757) | -0.37 | 0.588 |
| WBV | 186 | 1470 (1410, 1540) | 1460 (1400, 1520) | -0.74 | **0.002*** |
| EDSS | 181 | 2.5 (1.5, 5.0) | 3.0 (2.0, 6.0) | 20 | **<0.001*** |
| 9HPT | 47 | 22.3 (19.3, 25.6) | 23.7 (20.1, 27.6) | 6.5 | **<0.001*** |
| PASAT | 46 | 49.5 (37.2, 54.0) | 50.5 (42.0, 55.8) | 2.0 | 0.167 |
| SDMT | 49 | 53.0 (47.0, 64.0) | 51.0 (44.0, 59.0) | -3.8 | **0.010*** |
| T25FWT | 47 | 4.89 (4.27, 6.4) | 5.23 (4.58, 6.94) | 7.1 | **0.049** |

**Legend:** pwMS – people with multiple sclerosis, DGMV – deep gray matter volume, GMV – gray matter volume, LVV – lateral ventricular volume, WMV – white matter volume, WBV – whole brain volume, EDSS – Expanded Disability Status Scale, 9HPT – 9-Hole Peg Test, PASAT – Paced Auditory Serial Addition Test, SDMT – Symbol Digit Modalities Test, T25FWT – Timed 25-Foot Walk Test. Wilcoxon signed-rank test was used to compare between baseline and follow-up time points. P-values smaller than 0.05 were considered significant and are highlighted with bold fonts, and those with asterisks (*) survived the Benjamini-Hochberg correction for false discovery rate (FDR) correction.

### **Supplement Table 3.** Brain MRI scan volumes, EDSS, and neuropsychological test scores for patients with CIS/RRMS and PMS.

| **Time-point** | **Endpoint** | **CIS/RRMS median (IQR)** | **PMS median (IQR)** | **pwMS median (IQR)** | **P-Value** | **Age-Adjusted P-Value** |
| --- | --- | --- | --- | --- | --- | --- |
| **Baseline** | DGMV | 59.3 (55.8, 63.4) | 53.0 (49.3, 56.8) | 58.3 (53.0, 62.2) | **<0.001** | **0.001** |
|  | GMV | 753 (716, 789) | 711 (682, 735) | 742 (701, 781) | **<0.001** | 0.371 |
|  | LVV | 40.7 (32.0, 53.5) | 54.3 (39.3, 73.7) | 43.7 (33.9, 61.1) | **<0.001** | **0.033** |
|  | Thalamus | 19.8 (18.3, 21.2) | 17.4 (15.7, 19.0) | 19.3 (17.4, 20.8) | **<0.001** | **<0.001** |
|  | WMV | 732 (706, 766) | 687.0 (657.0, 724.0) | 723 (689, 752) | **<0.001** | **<0.001** |
|  | WBV | 1480 (1440, 1550) | 1390 (1360, 1460) | 1470 (1400, 1530) | **<0.001** | **<0.001** |
|  | EDSS | 1.5 (1.5, 2.5) | 6.0 (4.0, 6.5) | 2.5 (1.5, 4.88) | **<0.001** | **<0.001** |
|  | 9HPT | 21.3 (19.0, 24.1) | 27.6 (22.5, 41.5) | 22.3 (19.6, 25.6) | **<0.001** | **<0.001** |
|  | PASAT | 47.0 (37.0, 54.0) | 48.0 (26.0, 56.0) | 47.5 (36.0, 54.0) | 0.111 | 0.082 |
|  | SDMT | 55.0 (47.0, 65.0) | 47.5 (42.2, 50.8) | 52.0 (45.0, 62.5) | 0.021 | 0.062 |
|  | T25FWT | 4.84 (4.2, 6.17) | 6.76 (5.45, 9.71) | 5.16 (4.27, 6.98) | **<0.001** | **0.002** |
| **Follow-up** | DGMV | 57.1 (53.7, 60.8) | 50.7 (46.3, 55.0) | 55.7 (50.6, 59.2) | **<0.001** | **<0.001** |
|  | GMV | 755 (716, 787) | 705 (666, 735) | 739 (697, 775) | **<0.001** | **0.003** |
|  | LVV | 41.8 (32.9, 52.7) | 54.6 (39.6, 74.4) | 43.9 (35.1, 62.8) | **<0.001** | 0.123 |
|  | Thalamus | 19.1 (17.6, 20.4) | 17.0 (15.2, 18.0) | 18.4 (16.7, 19.8) | **<0.001** | **<0.001** |
|  | WMV | 734 (704, 767) | 693 (665, 730) | 722 (688, 757) | **<0.001** | **<0.001** |
|  | WBV | 1490 (1430, 1550) | 1400 (1330, 1450) | 1460 (1400, 1530) | **<0.001** | **<0.001** |
|  | EDSS | 2.0 (1.5, 3.0) | 6.5 (4.5, 6.5) | 3.0 (1.88, 6.0) | **<0.001** | **<0.001** |
|  | 9HPT | 21.4 (19.3, 23.7) | 28.6 (25.3, 37.0) | 23.4 (20.3, 29.3) | **<0.001** | **<0.001** |
|  | PASAT | 49.5 (41.8, 55.0) | 44.0 (31.0, 52.5) | 48.0 (37.0, 55.0) | **0.002** | **0.008** |
|  | SDMT | 56.0 (44.0, 63.0) | 44.0 (31.0, 48.2) | 49.5 (41.0, 59.0) | **<0.001** | **0.001** |
|  | T25FWT | 4.76 (4.3, 5.74) | 7.7 (6.33, 12.0) | 5.26 (4.45, 7.57) | **<0.001** | **<0.001** |

**Legend:** pwMS – people with multiple sclerosis, DGMV – deep gray matter volume, GMV – gray matter volume, LVV – lateral ventricular volume, WMV – white matter volume, WBV – whole brain volume, EDSS – Expanded Disability Status Scale, 9HPT – 9-Hole Peg Test, PASAT – Paced Auditory Serial Addition Test, SDMT – Symbol Digit Modalities Test, T25FWT – Timed 25-Foot Walk Test. T-test and Analysis of Covariance (ANCOVA) were used to assess the statistical significance of the difference between the two groups.

### **Supplement Table 4.** Changes in blood serum protein concentrations between baseline and the follow-up for all patients with proteome samples at both time-points.

| **Biomarker** | **Baseline median (IQR)** | **Follow-up median (IQR)** | **Percentage Change (%)** | **P-Value** |
| --- | --- | --- | --- | --- |
| **APLP1 (ng/mL)** | 12.0 (9.7, 14.6) | 12.7 (10.5, 14.5) | 5.7 | 0.239 |
| **CCL20 (pg/mL)** | 9.74 (6.01, 20.9) | 15.1 (8.01, 36.9) | 56 | **0.001*** |
| **CD6 (pg/mL)** | 139 (104, 189) | 142 (111, 181) | 2.0 | 0.867 |
| **CDCP1 (pg/mL)** | 107 (79.3, 140) | 108 (80.3, 158) | 1.3 | **0.008** |
| **CNTN2 (ng/mL)** | 1.85 (1.37, 2.49) | 1.95 (1.55, 2.76) | 5.3 | **0.016** |
| **CXCL13 (pg/mL)** | 50.4 (40.6, 69.8) | 51.5 (42.4, 69.6) | 2.2 | 0.555 |
| **CXCL9 (pg/mL)** | 54.4 (38.0, 88.5) | 64.8 (38.9, 99.4) | 19 | 0.089 |
| **FLRT2 (pg/mL)** | 114 (95.8, 137) | 118 (98.7, 135) | 3.0 | 0.562 |
| **GFAP (pg/mL)** | 122 (87.4, 164) | 129 (93.7, 175) | 5.4 | **0.041** |
| **IL-12B (pg/mL)** | 115 (77.8, 182) | 111 (78.2, 168) | -3.4 | 0.568 |
| **MOG (pg/mL)** | 28.4 (23.2, 35.9) | 31.3 (24.2, 37.7) | 10 | **0.020** |
| **NfL (pg/mL)** | 10.5 (7.75, 14.4) | 11.5 (8.57, 15.9) | 9.4 | **0.003*** |
| **OPG (pg/mL)** | 811 (658, 947) | 822 (668, 1050) | 1.4 | 0.089 |
| **OPN (ng/mL)** | 20.3 (15, 25) | 21.3 (16.6, 27.3) | 5.0 | **0.025** |
| **PRTG (pg/mL)** | 128 (110, 152) | 134 (113, 153) | 4.4 | 0.913 |
| **SERPINA9 (pg/mL)** | 52.7 (35.2, 81.5) | 54.9 (36.6, 77.8) | 4.2 | 0.931 |
| **TNFRSF10A (pg/mL)** | 6.01 (4.91, 7.51) | 6.41 (5.06, 7.8) | 6.6 | 0.145 |
| **TNFSF13B (ng/mL)** | 4.87 (4.17, 6.16) | 5.08 (4.19, 6.14) | 4.4 | 0.939 |
| **VCAN (pg/mL)** | 451 (388, 531) | 467 (392, 531) | 3.6 | 0.880 |

**Legend:** Wilcoxon signed-rank tested the significance in shifts. P-values smaller than 0.05 were considered significant and are highlighted with bold fonts, and those with asterisks (*) survived the Benjamini-Hochberg correction for false discovery rate (FDR).

### **Supplement Table 5.** Parameters of the linear model predicting the follow-up outcome score using single-protein models consisting of follow-up biomarker protein concentration, age, sex, BMI that passed the significance threshold of p=0.05.

| **Endpoint** | **Biomarker** | **Estimate** | **R-squared** | **P-value** |
| --- | --- | --- | --- | --- |
| DGMV | GFAP | -0.018 | 0.2 | **0.002*** |
|  | NfL | -0.013 | 0.17 | **0.021** |
|  | FLRT2 | -0.011 | 0.18 | **0.031** |
|  | TNFRSF10A | -0.006 | 0.33 | **0.029** |
|  | GFAP | -0.011 | 0.36 | **0.001*** |
| LVV | GFAP | 0.062 | 0.28 | **<0.001*** |
| Thalamus | GFAP | -0.015 | 0.19 | **0.009** |
|  | FLRT2 | -0.011 | 0.2 | **0.033** |
| WMV | GFAP | -0.006 | 0.15 | **0.018** |
|  | CXCL13 | -0.006 | 0.18 | **0.008** |
| WBV | GFAP | -0.009 | 0.33 | **0.001*** |
|  | CXCL13 | -0.005 | 0.3 | **0.022** |
|  | TNFRSF10A | -0.005 | 0.3 | **0.022** |
|  | NfL | -0.005 | 0.29 | **0.035** |
| EDSS | NfL | 0.054 | 0.31 | **0.002*** |
|  | GFAP | 0.043 | 0.26 | **0.021** |
|  | CXCL13 | 0.033 | 0.28 | **0.036** |
| 9HPT | VCAN | 0.026 | 0.18 | **0.04** |
|  | NfL | 0.046 | 0.24 | **0.001*** |
|  | GFAP | 0.043 | 0.21 | **0.003*** |
|  | FLRT2 | 0.034 | 0.2 | **0.01** |
| PASAT | NfL | -0.029 | 0.09 | **0.044** |
| SDMT | GFAP | -0.033 | 0.14 | **0.038** |
| T25FWT | NfL | 0.076 | 0.14 | **0.002*** |
|  | GFAP | 0.056 | 0.1 | **0.027** |

L**egend:** pwMS – people with multiple sclerosis, DGMV – deep gray matter volume, GMV – gray matter volume, LVV – lateral ventricular volume, WMV – white matter volume, WBV – whole brain volume, EDSS – Expanded Disability Status Scale, 9HPT – 9-hole peg test, PASAT – Paced Auditory Serial Addition Test, SDMT – Symbol Digit Modalities Test, T25FWT – Timed 25-foot walk test. P-values smaller than 0.05 were considered significant and are highlighted with bold fonts, and those with asterisks (*) survived the Benjamini-Hochberg correction for false discovery rate (FDR).

### **Supplement Table 6.** Parameters of a linear model predicting shift in outcome score using shift in biomarker concentration between baseline and follow-up along with age, sex, BMI that passed significant threshold.

| **Endpoint** | **Assay** | **Estimate** | **R-squared** | **P-value** |
| --- | --- | --- | --- | --- |
| **DGMV** | APLP1 | 0.004 | 0.04 | **0.022** |
|  | CD6 | -0.005 | 0.08 | **0.001*** |
|  | CDCP1 | -0.004 | 0.06 | **0.009** |
|  | CXCL9 | -0.004 | 0.05 | **0.018** |
|  | FLRT2 | -0.004 | 0.05 | **0.01** |
|  | GFAP | 0.003 | 0.03 | **0.038** |
|  | TNFRSF10A | -0.003 | 0.04 | **0.036** |
| **GMV** | TNFRSF10A | -0.004 | 0.04 | **0.035** |
| **Thalamus** | CDCP1 | -0.004 | 0.06 | **0.012** |
| **WMV** | GFAP | -0.007 | 0.07 | **0.005** |
|  | OPN | -0.005 | 0.05 | **0.025** |
|  | TNFSF13B | -0.005 | 0.05 | **0.037** |
| **WBV** | CDCP1 | -0.003 | 0.09 | **0.001*** |
|  | CNTN2 | -0.002 | 0.05 | **0.021** |
|  | CXCL9 | -0.002 | 0.04 | **0.048** |
|  | FLRT2 | -0.003 | 0.09 | **0.001*** |
|  | PRTG | -0.003 | 0.06 | **0.008*** |
|  | TNFRSF10A | -0.004 | 0.11 | **0.001*** |
|  | TNFSF13B | -0.004 | 0.10 | **0.001*** |
| **EDSS** | TNFSF13B | 0.022 | 0.08 | **0.021** |
| **9HPT** | CDCP1 | 0.019 | 0.18 | **0.040** |
| **PASAT** | NfL | -0.030 | 0.13 | **0.046** |
|  | OPN | -0.032 | 0.13 | **0.041** |
|  | VCAN | -0.030 | 0.12 | **0.039** |
| **SDMT** | NfL | -0.025 | 0.20 | **0.015** |
|  | PRTG | -0.023 | 0.18 | **0.029** |

**Legend:** pwMS – people with multiple sclerosis, DGMV – deep gray matter volume, GMV – gray matter volume, LVV – lateral ventricular volume, WMV – white matter volume, WBV – whole brain volume, EDSS – Expanded Disability Status Scale, 9HPT – 9-Hole Peg Test, PASAT – Paced Auditory Serial Addition Test, SDMT – Symbol Digit Modalities Test, T25FWT – Timed 25-Foot Walk Test. P-values smaller than 0.05 were considered significant and are highlighted with bold fonts, and those with asterisks (*) survived the Benjamini-Hochberg correction for false discovery rate (FDR).
